# Peripheral inflammation is associated with micro-structural and functional connectivity changes in depression-related brain networks

**DOI:** 10.1101/2020.09.09.20191262

**Authors:** Manfred G Kitzbichler, Athina R Aruldass, Gareth J Barker, Tobias C Wood, Nicholas G Dowell, Samuel A Hurley, John McLean, Marta Correia, Charlotte Clarke, Linda Pointon, Jonathan Cavanagh, Phil Cowen, Carmine Pariante, NIMA consortium, Mara Cercignani, Edward T Bullmore, Neil A Harrison

**Affiliations:** University of Cambridge, Brain Mapping Unit, Department of Psychiatry, Downing Site, Cambridge CB2 3EB, UK; Institute of Psychiatry, Psychology and Neuroscience, Department of Psychological Medicine, King’s College London, SE5 9RT, London, UK; University of Sussex, Brighton and Sussex Medical School, Clinical Imaging Sciences Centre, Brighton BN1 9RR, UK; Cardiff University Brain Research Imaging Centre, Cardiff University, Maindy Road, Cardiff, CF24 4HQ, UK; University of Oxford Department of Psychiatry, Warneford Hospital, Oxford, OX3 7JX, UK; University of Wisconsin, Department of Radiology, Madison, WI USA 53705-2275; College of MVLS, Institute of Health and Wellbeing, University of Glasgow, Glasgow, G51 4TF, UK; Centre for Immunobiology, University of Glasgow and Queen Elizabeth University Hospital, Glasgow, G51 4TF, UK; MRC Cognition and Brain Sciences Unit, Cambridge CB2 7EF, UK

**Author notes:** These authors contributed equally (Edward T Bullmore, Neil A Harrison).

**Keywords:** depression, inflammation, structural MRI, functional MRI

## Abstract

Peripheral inflammation can cause depressive symptoms – but how? Here, we measured brain MRI functional connectivity and micro-structural parameters, including proton density (PD, a measure of tissue water), at 360 cortical and 16 subcortical brain regions in 46 healthy controls and 83 depressed cases. Blood C-reactive protein (CRP) was positively correlated with PD in key precuneus/posterior cingulate cortex (pC/pCC) and medial prefrontal cortex (mPFC) components of the default mode network (DMN). CRP concentration was associated with increased connectivity within the DMN (nodes pC/pCC, mPFC and hippocampus), but decreased connectivity between DMN and non-DMN nodes. Depressed cases had reduced weighted degree, a metric of hubness, in anatomically co-located DMN regions. Our interpretation is that low-grade peripheral inflammation is associated with increased water (oedema) within a few highly connected but metabolically vulnerable medial cortical nodes, and this “rock in the pond” serves to perturb the functional connectivity of large-scale cortico-subcortical systems associated with depression.

## Introduction

Peripheral inflammation is strongly associated with depressive symptoms and behaviours. Studies in hepatitis patients therapeutically administered the pro-inflammatory cytokine interferon-α (Musselman et al., 2001), longitudinal epidemiological studies (Khandaker et al., 2018, 2014; Wium-Andersen et al., 2013; Liukkonen et al., 2006), investigations of co-morbid depression in systemic inflammatory disorders (Krueger et al., 2001; Tyring et al., 2006), and experimental studies in animal models (Dantzer et al., 2008), collectively provide compelling evidence that activation of the innate immune system is associated with increased risk of depression. They also provide strong evidence for the hypothesis that peripheral inflammation can *cause* depressive behaviours. However, the mechanistic chain of cause-and-effect linking increased peripheral inflammation to depressive mental states via intermediate effects on brain structure and function remains incompletely understood.

Some of the clearest evidence that peripheral inflammation can cause changes in human brain function has come from functional magnetic resonance imaging (fMRI) studies of brain activation in response to emotional or cognitive tasks. Some of these experiments have collected fMRI data before and after a planned inflammatory challenge, such as administration of IFN-α to hepatitis patients, or typhoid vaccine to healthy volunteers. These fMRI studies of experimentally manipulated inflammation are designed to test a causal relationship, and the pattern of results is consistent with observational studies that have reported correlations between task-related activation and experimentally uncontrolled between-subject variation in C-reactive protein (CRP) or other peripheral immune biomarkers. Meta-analysis of 24 primary studies (total N∼457) demonstrated replicable and significant effects of inflammation on brain activation in dorsal anterior cingulate cortex (dACC), subgenual anterior cingulate cortex (sgACC) and adjacent areas of medial prefrontal cortex (mPFC), insula, hippocampus, amygdala, and striatum (Kraynak et al., 2018).

Resting state fMRI studies have also investigated the relationship between peripheral inflammation and functional connectivity, usually measured by the correlation between a pair of regional fMRI time series. Seed-based correlational analysis, focused on a few *a priori* regions of interest, has shown that typhoid vaccination-induced increases in interleukin-6 (IL6, a pro-inflammatory cytokine) were negatively correlated with sgACC connectivity in healthy volunteers (Harrison et al., 2009); and CRP was negatively correlated with functional connectivity between striatal and amygdala subcortical regions and the mPFC in patients with major depressive disorder (MDD) (Felger et al., 2016; Mehta et al., 2018). Whole brain analysis of the relationship between fMRI connectivity and peripheral inflammation has produced more complex results. In healthy adults, both sgACC and dorsal mPFC demonstrated significant IL6-related changes in connectivity, but in opposite directions (Marsland et al., 2017). The human lipopolysaccharide (LPS) experimental model of inflammation-induced depression caused *increased* functional connectivity within thalamo-cerebellar and amygdalarfronto-parietal circuits (Labrenz et al., 2016); whereas the IFN-α model of inflammation-induced depression caused a global decrease in functional connectivity of sub-cortical (thalamus, striatum) and cortical areas (fronto-parietal, precuneus) (Dipasquale et al., 2016). In two samples of healthy african american participants (N=90 and N=82), increased peripheral inflammation (indexed by a composite of CRP, IL6, IL10 and TNF-α serum or plasma measurements) was associated with significantly reduced connectivity between components of an emotion regulation network (comprising medial premotor and lateral prefrontal, temporal and parietal cortices); but had no significant relationship with connectivity of the DMN (Nusslock et al., 2019).

These and other fMRI data broadly support the concept that there are “inflammation-sensitive” areas or networks of the human brain, many of which have also been reported to show depression-related differences in functional activation and/or connectivity in case-control fMRI studies of MDD (Fransson and Marrelec, 2008) (Marchetti et al., 2012; Menon, 2011). However, not all fMRI connectivity studies of inflammation have reported data for the hippocampus and other subcortical structures that have been implicated in the pathogenesis of depression (Schmaal et al., 2016) and have demonstrated inflammation-related changes in task activation (Marsland et al., 2008; Satizabal et al., 2012). A more fundamental limitation of fMRI as a marker of inflammation-related brain changes is its lack of cellular or molecular specificity.

In this context, micro-structural MRI is potentially interesting as a complementary approach to investigating the effects of inflammation on the human brain. Micro-structural MRI parameters describe the composition of tissue represented by each voxel, in contrast to more traditional macro-structural parameters, e.g., cortical thickness, which combine data from multiple voxels to measure brain anatomy. For example, magnetization transfer (MT) is a micro-structural MRI technique for measuring parameters which are biophysically interpretable in terms of the relative proportions of bound versus free water protons, or absolute proton density (PD). Studies in healthy adults have shown that typhoid vaccination and IFN-α administration both induced significant changes in MT parameters in the striatum and other structures (Dowell et al., 2016; Harrison et al., 2015) which were interpretable in terms of inflammation causing an increase in astrocyte-neuronal lactate shuttling and aerobic glycolysis (Pellerin and Magistretti, 1994).

Here we combined whole brain fMRI measurements of functional connectivity with MT measurements of micro-structure in an effort to elucidate how inflammation-related changes in the local, biophysical properties of brain tissue could be related to more distributed changes in functional connectivity between cortical and subcortical nodes of depression-related brain networks. Functional connectivity and micro-structural parameters were measured in the same set of 360 cortical areas and 16 subcortical regions in a sample of depressed cases (N=83, including 33 with CRP > 3 mg/L) and healthy controls (N=46). On this basis, we tested three principal hypotheses: (i) there are inflammation-related changes in quantitative MT parameters of brain tissue composition; (ii) there are inflammation-related changes in functional connectivity of cortico-subcortical networks; and (iii) inflammation-related changes in micro-structure and functional connectivity are anatomically co-located with each other and with depression-related changes in functional connectivity of cortico-subcortical networks.

## Results

### Sample

Socio-demographic and clinical data are summarised in **Table 1** for the sample with analysable fMRI data from 3 sites. Healthy controls were group mean-matched to all depressed cases on age and sex. As anticipated, there were significant case-control differences in depression, anxiety, fatigue, and anhedonia scores, as well as retrospectively self-reported childhood adversity exposure. Depressed cases with high CRP included a higher proportion of women (85%) and had higher BMI (29) than cases with low CRP (58% female, BMI = 26), as well as marginally lower childhood adversity scores (CTQ=49 vs 56). There were no significant differences between high and low CRP subgroups of cases on any other clinical variables.

**Table 1:** Socio-demographic and clinical data on the analysable sample from this case-control study of depression cases, stratified by high or low CRP, and healthy controls.

|  | Controls | Cases | p | loCRP Cases | hiCRP Cases | p |
| --- | --- | --- | --- | --- | --- | --- |
| n | 46 | 83 |  | 50 | 33 |  |
| sex (female/male) | 27/19 | 57/26 | 0.335 | 29/21 | 28/5 | 0.015 |
| age, years | 35.5 (7.5) | 37.1 (7.3) | 0.244 | 36.8 (7.1) | 37.6 (7.6) | 0.631 |
| body mass index | 24.5 (4.2) | 26.8 (4.1) | 0.002 | 25.6 (3.7) | 28.8 (3.9) | <0.001 |
| C-reactive protein, mg/L | 0.9 (0.7) | 2.9 (2.9) | <0.001 | 1.0 (0.7) | 5.8 (2.6) | <0.001 |
| clinician-rated depression<sup>a</sup> | 0.5 (0.9) | 19.3 (5.2) | <0.001 | 19.4 (5.4) | 19.2 (4.9) | 0.846 |
| self-rated depression<sup>b</sup> | 2.0 (2.8) | 25.3 (8.8) | <0.001 | 24.7 (8.4) | 26.2 (9.4) | 0.445 |
| state anxiety<sup>c</sup> | 26.9 (7.8) | 51.2 (10.7) | <0.001 | 51.2 (10.8) | 51.1 (10.7) | 0.957 |
| trait anxiety<sup>c</sup> | 29.4 (5.9) | 60.6 (9.2) | <0.001 | 60.3 (9.7) | 60.9 (8.6) | 0.767 |
| fatigue<sup>d</sup> | 10.7 (2.5) | 20.5 (5.7) | <0.001 | 20.4 (5.6) | 20.6 (5.8) | 0.866 |
| anhedonia<sup>e</sup> | 0.2 (0.6) | 5.0 (3.6) | <0.001 | 5.3 (3.4) | 4.7 (3.8) | 0.484 |
| childhood trauma<sup>f</sup> | 38.2 (5.1) | 53.2 (14.2) | <0.001 | 55.8 (14.9) | 49.2 (12.3) | 0.039 |
| Treatment Resistant | — | 50 | — | 28 (56%) | 22 (67%) | 0.367 |
| Number of antidepressants | — | 2.7 (1.7) | — | 2.6 (1.5) | 2.9 (2.0) | 0.405 |

<sup>a</sup> HAMD-17: Hamilton Rating Scale for Depression (Hamilton, 1960), a 17-item clinician-administered rating scale for reliable assessment of symptoms in patients diagnosed with depression (Kneisevich et al., 1977), with a total score range 0-52.
<sup>b</sup> BDI: Beck Depression Inventory version 2 (Beck and Beamesderfer, 1974), a 21-item self-report questionnaire, with a total score range 0-63.
<sup>c</sup> STAI: Spielberger State-Trait Anxiety Rating Scale (Spielberger, 1983), a 40-item self-report measure of trait and state anxiety.
<sup>d</sup> CFS: Chalder Fatigue Scale (Chalder et al., 1993), a 14-item self-report instrument to measure the severity of fatigue in adults.
<sup>e</sup> SHAPS: Snaith-Hamilton Pleasure Scale (Snaith et al., 1995), a 14-item self-report measure of anhedonia (loss of the normal capacity for pleasurable experience), one of two principal symptoms of depression.
<sup>f</sup> CTQ: Childhood Trauma Questionnaire (Bernstein et al., 1994), a standardized, retrospective 28-item self-report inventory that measures the severity of five classes of childhood trauma: emotional abuse, physical abuse, sexual abuse, emotional neglect, and physical neglect.

MT scans were excluded from one assessment centre (KCL) because pilot studies indicated insufficiently close alignment between the data collected at this site compared to the other two assessment centres, which used identical 3T MRI systems. Consequently, the sample size for micro-structural analysis is smaller (**Table S2**) than for fMRI analysis **Table 1**.

### Proton density: between-group differences and correlation with CRP

We estimated the global brain density of each qMT parameter, across all 360 cortical and 16 sub-cortical regions, on average for controls, all cases and each sub-group of cases. This demonstrated some significant between-(sub)group differences in the distributions of PD (Kolmogorov Smirnov (KS) tests, P < 2.1×10^−6^; **Figure 1A**). The greatest distributional difference in PD was between high CRP (≥ 3 mg/L) and low CRP (< 3 mg/L) subgroups of cases (P < 6.4×10^−7^); and the smallest distributional difference was between low CRP cases and controls (P < 0.72). Between-group differences were not significant for the global distributions of most other qMT parameters (k_bf_, f_b_, T2_b_, T1_f_; see SI **Figure S2**); although there was a nominally significant between-group difference in the distribution of the T2_f_ parameter (transverse relaxation time of the free component; KS test P < 0.0005).

**Figure 1:**
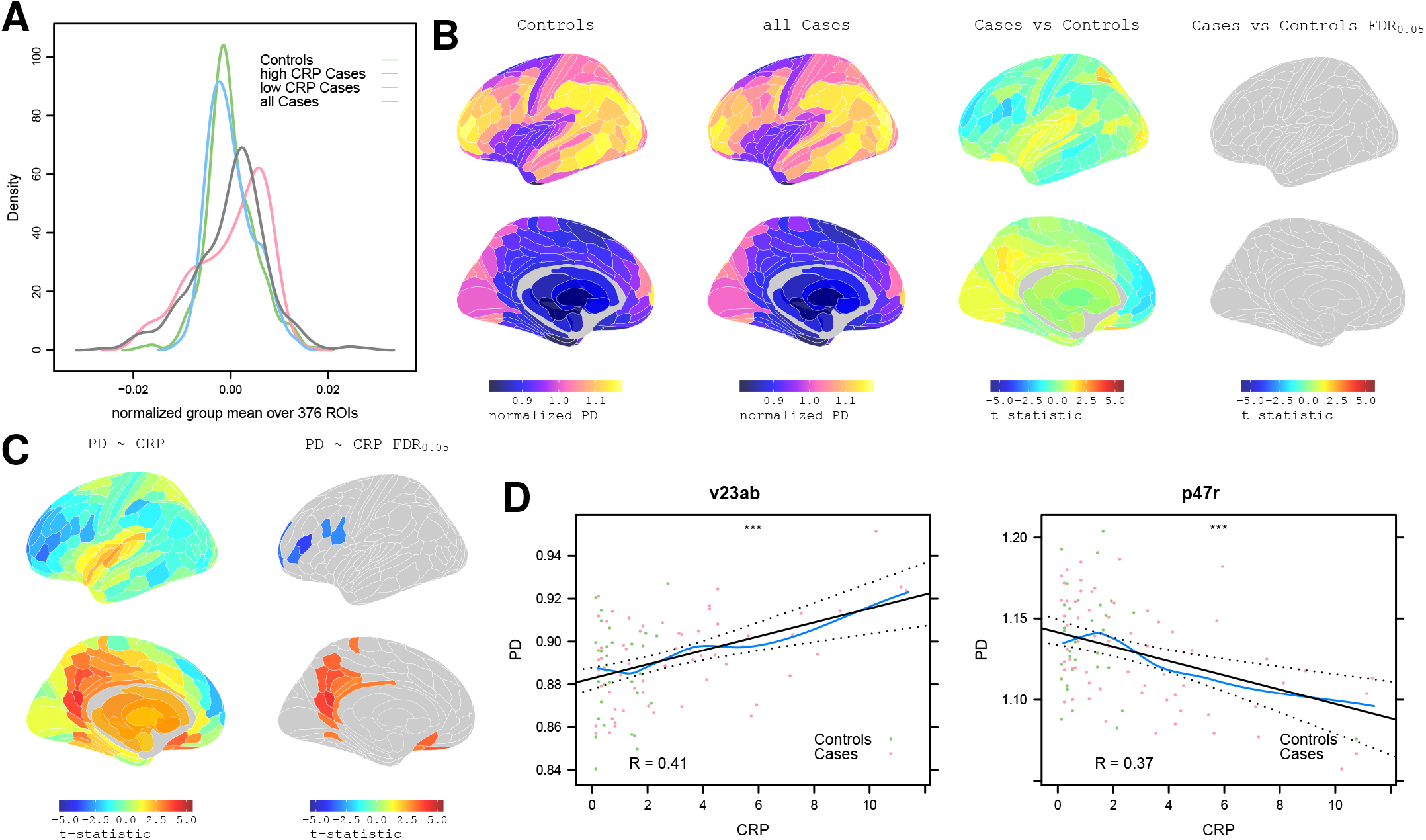
**Proton density MRI: between-group differences and correlation with CRP**. (a) Densities of normalized group mean PD over 376 regions for all cases, all controls, and for high CRP and low CRP subgroups of cases. (b) From left to right, cortical maps of: PD for all controls, PD for all cases, case-control difference in PD (*t*-statistic), and significant case-control differences in PD with regional P_FDR_ < 0.05. (c) Left, cortical *t*-statistic map of association between regional PD and blood CRP concentration, *PD* ∼ *CRP*, in all cases and controls combined; right, map of regions where *PD* ∼ *CRP* was significantly greater than zero (orange regions), or less than zero (blue regions), with regional P_FDR_ < 0.05. (d) Left, scatterplot of PD (y-axis) versus CRP (x-axis) for posterior cingulate cortical region v23ab; right, scatterplot of PD versus CRP for dorsolateral prefrontal cortical region, p47r.

We estimated and tested depression- and inflammation-related effects on PD at each of 376 brain regions. The resulting map of the *t*-statistic for the all cases-controls difference in PD demonstrated small effect sizes, none of which was significantly greater than expected under the null hypothesis after controlling for multiple comparisons (P_FDR_ < 0.05) (**Figure 1B**). Taken together with the convergent distributional results in global PD, these regional data indicated that there was not a major effect of depression on PD. Therefore, we proceeded to investigate the association between PD and CRP using data from all participants (cases and controls combined).

We regressed PD on CRP at each region, resulting in a parcellated map of the association between PD and CRP (**Figure 1C-D**), denoted *PD* ∼ *CRP*, with localised strong positive and negative correlations that were statistically significant (P_FDR_ < 0.05) in 22 regions. Posterior cingulate cortex and precuneus (pC/PC; 10 regions) - RSC, PCV, 7m, POS1, v23ab, d23ab, 31pv, 31pd, 31a, ProS; inferior, orbital and polar frontal cortex (6 regions) - IFJa, IFSa, a10p, p10p, p47r, OFC; anterior cingulate and medial prefrontal cortex (2 regions) - pOFC, 25; and single regions of dorsolateral prefrontal, premotor, paracentral and ventral visual stream cortex - 9a, 6r, 5m, VMV1. Negative correlations with CRP were concentrated in 7 regions of prefrontal and premotor cortex; positive correlations were located in 15 regions of posterior and anterior cingulate cortex (see **Table S3**).

### Functional connectivity: between-group differences

We plotted the distributions of all pair-wise fMRI time series correlations or edge weights (70,500) on average over all participants in each (sub)group (**Figure 2A**, left). These distributions were generally positive, but differed significantly between cases and controls (Kolmogorov-Smirnov test, P < 2.2×10^−16^). Specifically, depressed cases had a functional connectivity distribution shifted to the left compared to controls, indicating a greater proportion of negatively weighted edges, especially in the high CRP subgroup.

**Figure 2:**
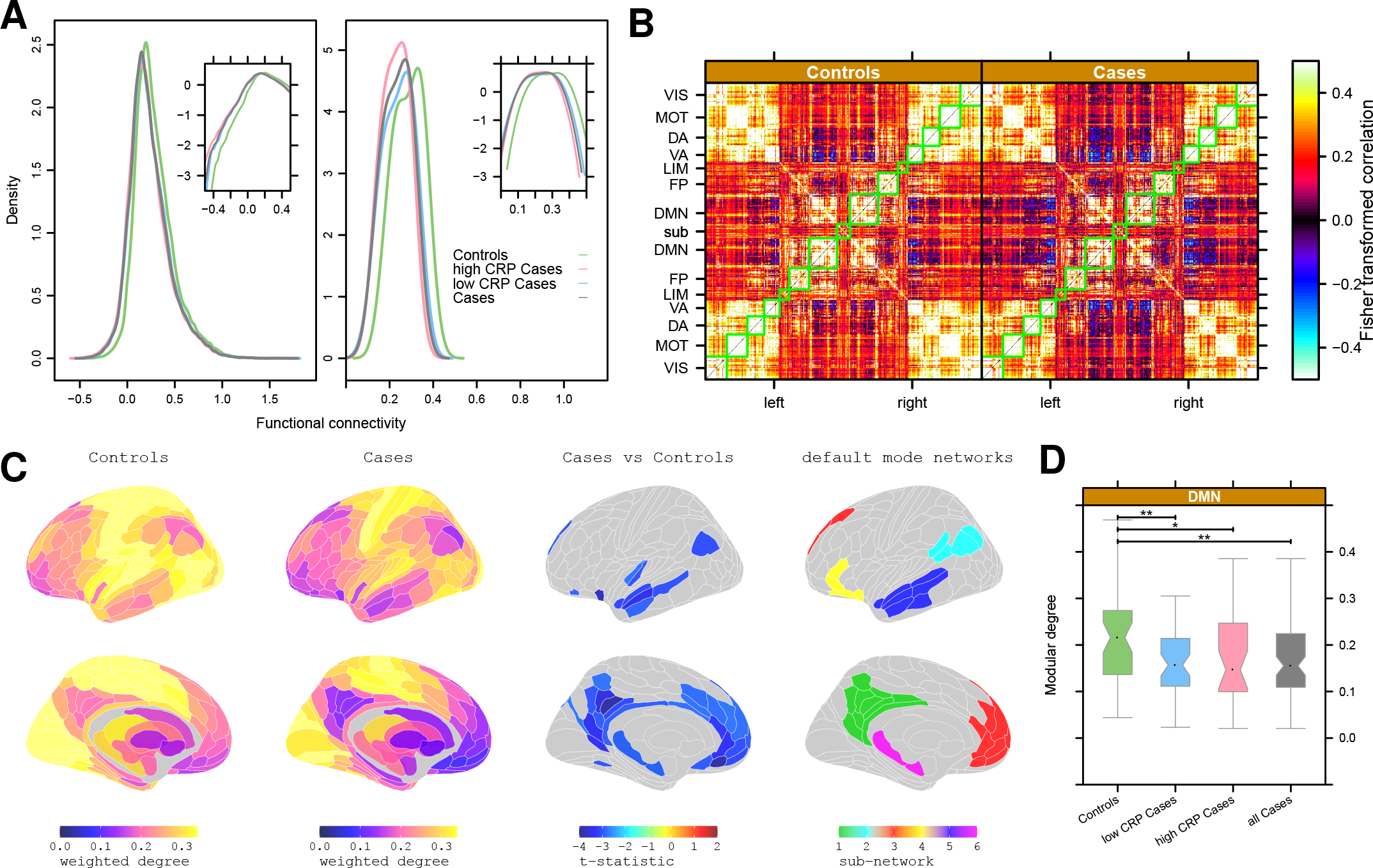
**Functional connectivity: between-group differences**. (a) Left, group mean density of pair-wise correlations or edge weights for all controls, all cases, and high or low CRP subgroups of cases. Right, group mean density of weighted degree or nodal hubness for all controls, all cases, and high or low CRP subgroups of cases. Insets show the same data on a log scale to clarify between-group differences in the negative (or less positive) tails of the distributions. (b) Group mean inter-regional correlation or functional connectivity matrices for all controls (left) and all cases (right). Each row or column in these symmetrical matrices represents one of 360 cortical regions which have been ordered by their prior affiliation to resting state networks or modules: VIS = visual, MOT = motor, DA = dorsal attentional, VA = ventral attentional, LIM = limbic, FP = fronto-parietal, DMN = default mode network. 16 subcortical regions are designated SUB. (c) Cortical and subcortical maps of group mean weighted degree for controls and cases, and the map of significant case-control differences in weighted degree (P_FDR_ < 0.05), demonstrate reduced hubness of nodes in many cortical areas of the default mode network (DMN, right column). (d) Boxplots of weighted degree of all cortical nodes in the DMN for all controls, all cases, and high or low CRP subgroups of cases. ** denotes significant between-group differences in modular degree of DMN (P < 0.05).

To localise these between-group differences in functional connectivity, we estimated the mean correlation matrix for each group and organised the regional rows and columns in alignment with a prior set of resting state networks or modules (Yeo et al., 2011) **Figure 2B**. There was a clear block-diagonal pattern for both the case and control matrices, indicating strong positive correlations between nodes within the same module, especially the visual, motor, dorsal attentional (DA) and ventral attentional (VA) modules. Off-diagonal correlations between modules were generally weaker, although strong positive correlations between visual, motor, DA and VA modules were evident. In contrast, there were strong negative correlations between the default mode network (DMN) and the VA and DA networks. The modular community structure of brain functional networks was qualitatively similar between groups; but depressed cases had stronger negative correlations between the DMN and other modules compared to controls.

The distribution of group mean weighted degree of functional connectivity was estimated over all regional nodes for cases and controls (**Figure 2A**, right). Weighted degree was always positive in the range 0.07-0.42. In depressed cases, the degree distribution was significantly shifted to the left, indicating fewer nodes with high degree and more nodes with near-zero degree (KS statistics: for controls vs cases, and controls vs high CRP cases, P < 2.2×10^−16^; for controls v low CRP cases P < 1.8×10^−13^; for low CRP vs high CRP cases, P < 5×10^−5^).

To localise these global differences, mean weighted degree was represented as a cortical map for each group (**Figure 2C**).

Hub nodes, with high weighted degree, were concentrated in somatosensorimotor, visual and auditory cortices; non-hub nodes, with low degree, were concentrated in DMN cortical areas and sub-cortical structures. Negative case-control differences in weighted degree, indicating reduced nodal connectivity in depressed cases, were statistically significant (P_FDR_ < 0.05) in 39 regions of pC/pCC, inferior parietal cortex, mPFC and hippocampus (see SI **Table S3**). Many of these regions of significantly reduced hubness in depressed cases were affiliated to the default mode network (**Figure 2C**). More formally, the group mean modular degree of all nodes within each of the prior modules was specifically and significantly reduced for depressed cases compared to controls in the DMN (**Figure 2D**), but not in any other cortical modules (**Figure S3**)

### Functional connectivity: correlation with CRP

In light of the significant depression-related differences in functional connectivity, we estimated the association between CRP and functional connectivity using only data from depressed cases (not including controls). We first regressed CRP on each of 70,500 inter-regional correlations, and on each of 376 nodal degrees. This identified four statistically significant connections that survived FDR correction at P < 0.05. Notably, three of these were between DMN nodes including connections between the pC/pCC (areas RSC, v23ab, POS1) and the hippocampus, and between the hippocampus and mPFC (area 10r), both of which showed a positive correlation with CRP; a connection between an area abutting the pC/pCC (area 7Pm) and the frontal-opercular area (FOP1) showed a significant negative correlation with CRP (**Figure 3A**)

**Figure 3:**
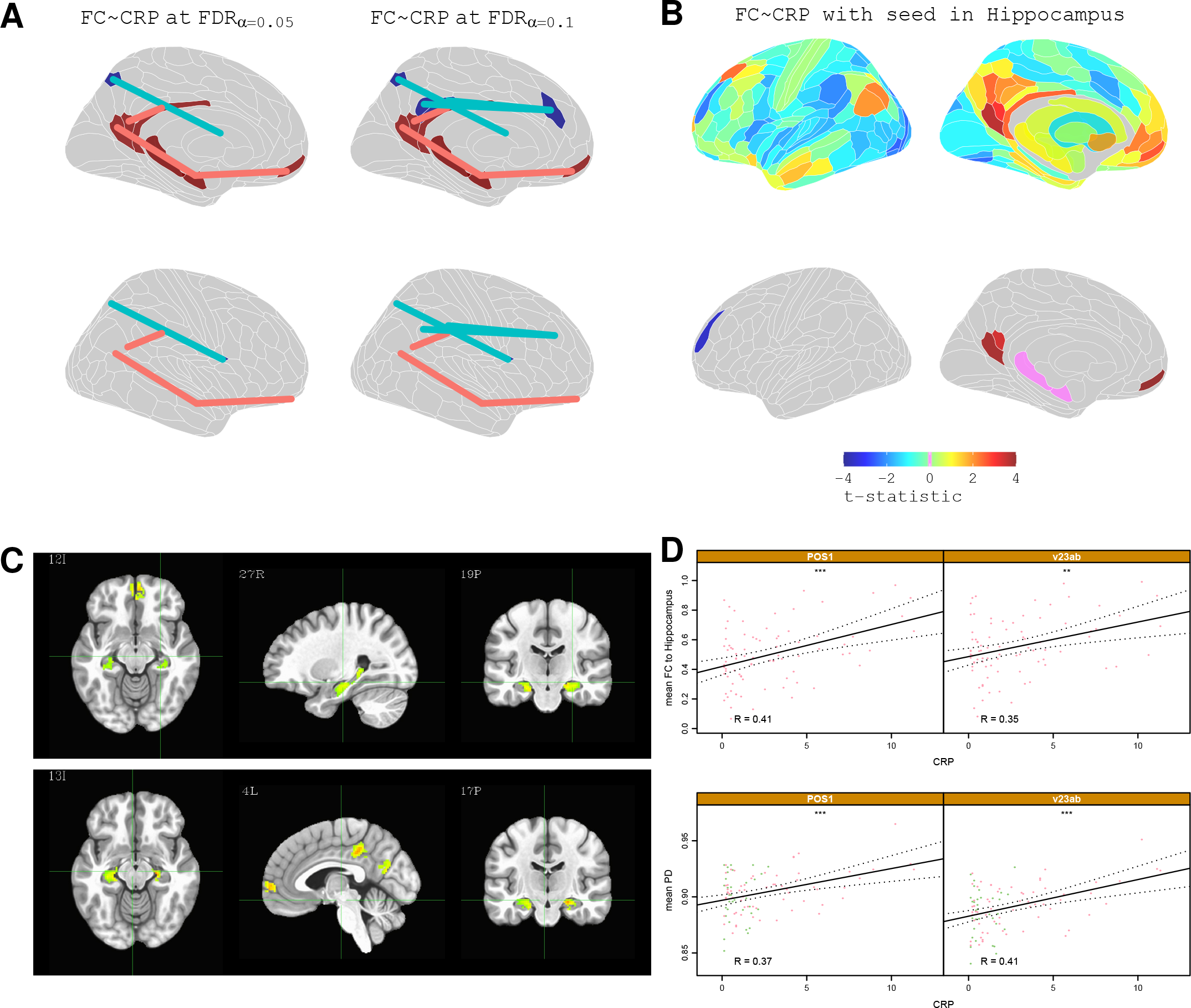
**Functional connectivity: correlation with CRP**. (a) Cortical maps highlighting edges between regional nodes defined by significant correlation of functional connectivity with CRP (FDR corrected at 0.05 and 0.1) (b) Top row, brain maps of *t*-statistics representing strength of association between CRP and functional connectivity of the hippocampus (with each one of all other brain regions). Bottom row, cortical maps highlighting regions where CRP was significantly, positively correlated with functional connectivity with the hippocampus (P_FDR_ < 0.05). (c) Voxel based analysis of functional connectivity dependence on CRP for a seed in the PCC (top row) and for a seed in the right Hippocampus (bottom row). Functional connectivity between hippocampus, PCC, and mPFC was positively correlated with CRP (only significant clusters with α < 0.05 are shown; global significance threshold P < 0.05). (d) Scatterplots illustrating the relationship between CRP and hippocampus connectivity for two cortical regions: POS1 and v23ab. Bottom, scatterplots of CRP vs PD for the same cortical regions; * denotes P < 0.05.

To investigate this in more detail, we used seed-based analyses to test the strength of association between CRP and the functional connectivity of each subcortical seed region with each one of all 360 cortical regions. This analysis showed that high CRP was associated with increased functional connectivity between the hippocampus and 6 regions predominantly in the pC/pCC and mPFC (**Figure 3B-C**; see **Table S4** for details). Conversely, high CRP was associated with reduced functional connectivity between the putamen and 9 regions of DLPFC, pC/pCC, inferior and superior parietal cortex (see **Table S4** for details), and reduced functional connectivity between thalamus and a set of four, partly overlapping regions of DLPFC and inferior parietal cortex (see **Table S4** for details). Notably, many of these areas also showed scaling between PD and CRP: namely, pC/pCC areas POS1 and v23ab showed significantly increased functional connectivity to the hippocampus, as well as significantly increased PD, in association with high CRP (**Figure 3D**); and pC/pCC areas 7m and d23ab showed significantly reduced functional connectivity to the putamen, as significantly increased PD. One frontal region, 6r, that had significantly decreased PD in participants with high CRP also showed significantly reduced connectivity to the thalamus. Together, these findings suggest a pattern in which CRP positively scales with the functional connectivity of subcortical structures (i.e. hippocampus) affiliated to the DMN but negatively with cortical regions outside the DMN.

## Discussion

This study was designed to test three principal hypotheses. First, that there are peripheral inflammation-related changes in human brain micro-structure. Second, that there are inflammation-related changes in functional connectivity of cortico-subcortical networks. And third, that inflammation-related changes in brain micro-structure and functional connectivity are co-located with each other and with depression-related changes in functional networks. Our results provide new evidence in support of all three hypotheses.

### Inflammation-related changes in brain micro-structure

The clearest signal of inflammation-related change in the human brain was provided by the micro-structural measurements of proton density (PD). This was evident at a global scale as a significant difference in whole brain distributions of PD between high and low-CRP depressed cases (**Figure 1A**). It could also be resolved at the regional scale in terms of a set of 22 cortical regions in which PD was significantly correlated with CRP (**Figure 1C**). These inflammation-sensitive areas were restricted to higher order association cortices, particularly within the default mode network. In pC/pCC and mPFC components of the default mode, PD showed a positive scaling with CRP. However, in the DLPFC an inverse correlation between PD and CRP was observed. Notably, these key results were robust to several sensitivity analyses including statistical correction for potentially confounding variables and subgroup analyses restricted to depressed cases only (see **Figure S4**).

Proton density measures the concentration of MRI-visible protons at each voxel of the image. In the brain, the majority of protons are in water, and PD is widely interpreted as a measurement of tissue-free water content (Cercignani et al., 2018). Although all 1H MR image contrasts are influenced to some extent by PD, bespoke sequences such as used here to provide a precise quantitative measure of PD, are infrequently used in clinical research. Exceptions include studies of stroke, glioma, and multiple sclerosis, pathologies that all include a marked central inflammatory response, (Raschke et al., 2019; Kikinis et al., 1999) where increases in PD are commonly interpreted as a marker of inflammation-induced extracellular fluid accumulation, i.e., oedema. In this context, the simplest interpretation of our findings is that low grade peripheral inflammation (regardless of depression status), is associated with localised increases in extracellular fluid within pC/pCC and mPFC components of the default mode network.

Localized, peri-lesional increases in PD are characteristic of inflammatory responses to brain tumors, MS lesions and stroke (Raschke et al., 2019; Warach et al., 1995). There is also prior evidence that brain regions are differentially sensitive to systemic inflammation. For example, a clinical study of Aicardie-Goutier’s syndrome (Uggetti et al., 2009) and experimental-medicine studies of patients receiving interferon-alpha therapeutically (Dowell et al., 2016; Haroon et al., 2014) both illustrate the sensitivity of basal ganglia micro-structure and neurochemistry to chronically elevated type I interferons. Such studies have also demonstrated selective impairments in the functional connectivity of specific corticostriatothalamic circuits comprising higher order association cortical areas such as the pC/pCC and other components of the DMN (Dipasquale et al., 2016; Harrison, 2016). Covergently, animals experiments have demonstrated the selective recruitment of circulating monocytes to stress-responsive brain regions such as the mPFC and hippocampus (D’Mello et al., 2009; Wohleb et al., 2015).

In addition to this prior support for inflammation-responsive brain regions, there is some evidence more directly indicating that peripheral inflammation can cause changes in brain water content that are measurable by PD. For example, in rodents, peripheral endotoxin challenge resulted in an increase in cortical (but not cerebellar) tissue water content measured using a sensitive gravimetric technique (Wright et al., 2007). In humans, studies of hepatic encephalopathy, a peripherally mediated disorder believed to have an inflammatory etiology, reveal heterogenous increases in grey matter PD, particularly within the basal ganglia (Shah et al., 2008), and associated disturbances in functional connectivity (Qi et al., 2012; Jao et al., 2015) which have been interpreted as resulting from impaired connectivity of oedematous areas in cortical-basal ganglia loops.

Why these highly connected default mode regions are so susceptible to peripheral inflammation cannot be directly addressed in the current study, although it may be related to their high metabolic demands. For example, the pC/pCC has the highest cortical glucose metabolism (Gusnard and Raichle, 2001) and the highest expression of the mitochondrial oxidative phosphorylation enzyme cytochrome c oxidase in the brain (Vogt and Laureys, 2005). It also demonstrates the greatest reduction in cortical blood flow during sleep. However, there are some aspects of our data which are somewhat anomalous in relation to the interpretation that inflammation causes extracellular oedema measurable by increased PD. For example, PD was negatively correlated with CRP in a few areas of lateral prefrontal cortex, implying reduced free water concentration in association with peripheral inflammation. It is also notable that although we measured several other micro-structural MRI parameters, including T1, that have previously been interpreted as markers of free water concentration in white and grey matter (Mezer et al., 2013), only PD was robustly correlated with CRP in this study of cortical and subcortical grey matter.

### Depression-related changes in functional connectivity

The clearest signal of depression-related change was provided by the resting state fMRI measurements of connectivity between nodes, and the weighted degree or hubness of nodes, in the functional connectome. This was evident at a global scale by significant between-group differences in the distribution of edge weights and nodal degree. Depressed cases had more strongly negative edges and reduced degree or hubness of nodes (**Figure 2A,B**). This depression-related effect on functional connectivity could also be resolved at the regional scale as a set of 39 cortical areas and one subcortical (hippocampus) area whose weighted degree was significantly reduced in depressed patients compared to controls (see **Figure 2C** and **Table S3**). Notably, 27 (69%) of the regions with reduced nodal degree were affiliated to the default mode network. These key results were robust to several sensitivity analyses including statistical correction for BMI, head movement and other potentially confounding variables (see **Figure S4**).

Dysfunction within nodes of the default mode network (DMN) was among the first reported findings in depression (Drevets et al., 1997) and subsequent studies have reported altered resting state functional connectivity across each of its components (Zhu et al., 2012). Collectively, these studies reveal a pattern of *increased* functional connectivity between nodes intrinsic to the DMN (the so called task negative network) but reduced connectivity of DMN nodes to wider areas outside of the DMN (task postive networks) in depression (Marchetti et al., 2012; Menon, 2011). Within the DMN, interactions of the pC/pCC with virtually all other DMN nodes indicate a pivotal role for this posterior medial cortical area in regulating the flow of intrinsic activity throughout the network (Fransson and Marrelec, 2008). Furthermore, meta-analyses of seed-based studies have particularly highlighted the importance of dysregulated connectivity between the pC/pCC and the ventral attention and frontoparietal networks for the emergence of ruminative thoughts and emotional dysregulation that are characteristic clinical features of depression (Kaiser et al., 2015). Our finding of reduced nodal degree within 39 areas largely restricted to the DMN expands these findings to reveal a broader depression-related disconnection of the DMN from wider task positive areas.

### Inflammation-related changes in functional connectivity

Our analysis of significant scaling between CRP and functional connectivity identified four (out of 70,500) edges which survived stringent correction for multiple comparisons. Notably three of these edges mediated connections within the DMN (between hippocampus and pC/pCC (v23ab), and between hippocampus and medial prefrontal cortex (10r)) which were stronger in association with high CRP. Connectivity of a further area abutting the pC/pCC (area 7Pm) to the frontal-opercular area (FOP1) showed a negative scalling with CRP. More restricted analysis, focusing on the 360 edges connecting each subcortical structure to each of all cortical area confirmed the positive scaling of CRP with functional connectivity of the hippocampus to cortical areas within the DMN. It also identified significant CRP-related attenuation of functional connectivity between the putamen and a set of 9 cortical areas, of which four (44%) were affiliated to the DMN, and attenuated connectivity of the thalamus to a largely overlapping set of cortical regions. Together, this pattern of findings suggests that higher CRP is associated with increased functional connectivity of cortico-subcortical connections within the DMN but a decrease in DMN connectivity to subcortical regions outside of the DMN. Again, these key results were robust to several sensitivity analyses including statistical correction for BMI and other potentially confounding variables (see **Figure S4**).

### Implications for inflammation-associated depression

What are the implications for our motivating pathogenic hypothesis that peripheral inflammation predisposes to depression by proximal effects on brain structure and function? One formulation compatible with these data is that highly connected and metabolically active regions such as the pC/pCC are particularly susceptible to peripheral inflammation which causes a localised increase in extracellular water (indexed by PD). The pC/pCC is a critical hub within the DMN, and central to both the regulation of information flow throughout the DMN (via connections to the hippocampus and mPFC), and supression of DMN activity during task performance (Fransson and Marrelec, 2008). Functional connectivity of the pCC, mPFC and hippocampus have also been repeatedly implicated in the aetiology of depression (Marchetti et al., 2012) and demonstrated to be disrupted in both our current data and in meta-analysis of seed-based connectivity studies (Kaiser et al., 2015).

Crucially, in these data, regions of pC/pCC showing CRP-related changes in micro-structure (PD) also showed a concomitant CRP-dependent increase in within DMN connectivity (to hippocampus and consequently mPFC).

Together, these findings are consistent with a mechanistic model by which locally restricted effects of peripheral inflammation on brain micro-structure (oedema) can perturb the functional connectivity between inflammation-responsive regions and a more extensive cortico-subcortical network, which in turn gives rise to changes in emotion and cognition that are diagnosed clinically as depression. In other words, although only a subset of DMN regions are structurally sensitive to disruption by peripheral inflammation this localised inflammatory shock can propagate more widely by functional dysconnectivity throughout the DMN to result in depression (Marchetti et al., 2012). This formulation is compatible with the anatomical overlap between the relatively circumscribed inflammation-related changes in PD and functional connectivity and the more extensive depression-related changes in functional connectivity that we have demonstrated.

Further investigation of this hypothesis could benefit from animal experiments, measuring the effects of systemic inflammatory stimuli on PD and functional connectivity measured by MRI and on brain water concentration measured directly and near-simultaneously in tissue. Animal experiments will likely also be important in elucidating more precisely the mechanisms by which peripheral inflammation causes localised brain oedema and how this disrupts function. We have used CRP as a convenenient and well-established proxy for peripheral innate immune system activation; but we don’t assume that CRP itself is moving across the blood brain barrier (BBB) to cause local inflammatory changes in the brain. It seems more likely that high CRP is a surrogate for increased blood concentrations of pro-inflammatory cytokines or myeloid immune cells which can traffic across the BBB to have inflammatory effects on some brain regions.

### Methodological issues

There are some important limitations of this study to consider. The sample size is not large so there is an appreciable risk of inadequate power to detect small effects of depression or inflammation, especially in the context of stringent *P*-value thresholds adopted to correct for multiple comparisons. The depressed cases were enriched for patients with CRP greater than 3 mg/L, leading by design to a case-control difference in inflammation, or potential confounding of depression- and inflammation-related effects on MRI markers, although this risk was mitigated by the lack of evidence for depression-related changes in PD and restricting our analysis of CRP-related changes in functional connectivity to data acquired from cases only. Case-control designs are also vulnerable to the confounding effects of other variables on MRI biomarkers. In our principal analysis, we statistically controlled for age and scanning centre, on the grounds that they could affect the MRI metrics and were randomly different to a degree between groups. We did not principally control for effects of sex, BMI and experience of childhood adversity, given the prior evidence that all of these factors are known risk factors for both depression and inflammation. However, we demonstrated by sensitivity analyses that our key results were conserved after statistical correction for BMI and childhood adversity, and in analysis of females only. Head movement during scanning is a well-recognised source of bias in functional MRI connectivity analysis. Our principal analysis was predicated on a well-validated pre-processing pipeline for movement correction and all data passed standard quality control criteria for head movement before statistical analysis. We also included Global Signal Regression (Power et al., 2018) as one of the alternative methods in the sensitivity analysis (**Figure S4**) demonstrating that qualitatively the results did not change.

### Conclusions

Peripheral inflammation was associated with micro-structural changes in proton density, compatible with localised oedema of inflammation-responsive brain regions in the DMN, and with changes in functional connectivity of DMN areas, especially the hippocampus. These inflammation-related changes in micro-structure and functional connectivity were concentrated in areas of a default mode cortico-subcortical network that also demonstrated reduced functional connectivity in depression. These results help to bridge the mechanistic gap between peripheral inflammation and depression by demonstrating intermediate changes in human brain structure and function.

## Data Availability

Data and processing scripts will be made available.

## Funding acknowledgements

This study was funded by an award from the Wellcome Trust (grant number: 104025/Z/14/Z) for the Neuroimmunology of Mood Disorders and Alzheimer’s Disease (NIMA) consortium, which was also funded by Janssen, GlaxoSmithKline, Lundbeck and Pfizer. Additional support was provided by the NIHR Cambridge Biomedical Research Centre. ETB is an NIHR Senior Investigator.

## Author Contributions

GJB, TCW, SH, JM, MC collected the data. MGK, ARA, CC and NGD analysed the data. MGK, ETB, and NAH wrote the paper.

## Material and Methods

### Study design and sample

This was an observational, multi-site, case-control study. Depressed cases screened positive for current depressive symptoms on the Structured Clinical Interview for DSM-5 (SCID; First et al., 2016) and had total score greater than 13 on the Hamilton Rating Scale for Depression (HAM-D; Hamilton, 1960). Healthy controls screened negative for past or current depressive disorder on the SCID. All participants satisfied additional inclusion criteria, e.g. aged 25-50 years, and exclusion criteria, e.g. major medical inflammatory disorder or immuno-modulatory medication, as detailed in Supplementary Information (SI) **Table S1**. Depressed cases were stratified by venous blood concentration of CRP: high CRP cases (N=33) had CRP > 3 mg/L; low CRP cases (N=50 had CRP < 3 mg/L. All controls had CRP < 3 mg/L.

After preliminary telephone screening, potentially eligible participants attended one of 5 UK recruitment centres (Brighton, Cambridge, Glasgow, King’s College London (KCL), or Oxford) for clinical interviews and blood sampling for CRP. Eligible participants then attended one of three UK assessment centres (Cambridge, KCL, Oxford) for venous blood sampling, clinical assessment, and a single MRI scanning session; see SI **Figure S1** for details. Between-site reliability of fMRI connectivity and MT parameters was established by a pilot study of 5 healthy volunteers, each scanned at all three assessment centres, confirming that the coefficient of variability (CoV) was lower between sites than between subjects.

We collected complete data from 143 eligible participants categorised into three groups: healthy controls (HC, N=46), depressed cases with CRP < 3 mg/L (loCRP MDD, N=53), and depressed cases with CRP > 3 mg/L (hiCRP MDD, N=34). All groups were matched for mean age, sex and handedness (Edinburgh Handedness Inventory). Subject to quality control criteria applied to MRI and other data, the final, evaluable dataset is summarised in Results, **Table 1** and SI **Figure S1**.

All procedures were approved by an independent national research ethics service (NRES) committee (NRES: East of England, Cambridge Central, UK; Reference: 15/EE/0092). All participants provided written informed consent and received up to £325 reimbursement.

### Additional clinical assessments

All participants additionally completed the following self-report standardized instruments (see also footnotes **Table 1** and SI): Beck Depression Inventory version 2 (BDI-II), Snaith-Hamilton Pleasure Scale (SHAPS), State-Trait Anxiety Inventory (STAI), Chalder Fatigue Score (CFS), Childhood Trauma Questionnaire (CTQ), Perceived Stress Scale (PSS) and Life Events Questionnaire (LEQ).

### Biomarker assessments

We collected 50 mL of venous blood at 8-10 a.m. on the day of assessment. Participants had fasted since 10 p.m. the previous night and had been lying supine for 30 mins prior to venepuncture. C-reactive protein was measured using a high sensitivity assay at a single central laboratory (Q^2^ Solutions, The Alba Campus, Livingston EH54 7EG, UK) from 0.5 mL of plasma (reportable range 0.2-9999.9 mg/L).

Body mass index (BMI) was measured as weight (kg) divided by height squared (m^2^).

### Structural MRI data acquisition and pre-processing

Quantitative magnetization transfer (qMT) images were acquired using a magnetization transfer-weighted spoiled gradient echo sequence comprising a set of 10 combinations of flip angle (2: 360 and 720 deg) and off-resonance frequency (5: 1-25 kHz) with the following parameters: acquisition time = 20 min; relaxation time (TR) = 32 ms; echo time (TE) = 2.9 ms; field of view (FoV) 192 mm, equivalent to 64 axial slices at matrix size 80×80 and voxel size 2.4×2.4×2.5 mm.

The qMT data were realigned to subject specific structural images using rigid-body registration; qMT parameters were then estimated by voxel-wise non-linear least squares fitting (Levenberg-Marquardt) of a binary spin bath model using the QUIT package (QUantitative Imaging Tools; Wood, 2018). This yielded whole-brain maps of proton density (PD), bound proton fraction (f_b_), MT exchange rate (k_bf_), and the transverse relaxation times of the bound and free water components (T2_b_, T2_f_). Spatial intensity variations (bias field, RF inhomogeneities) were corrected using FSL FAST (Zhang et al., 2001). In order to account for differences in overall scanner sensitivity, we divided regional PD values by mean PD per subject, resulting in PD measurements globally normalized to unity.

Quantitative MT parameter maps were then regionally parcellated into 360 cortical regions, defined *a priori* using a well-validated parcellation template (Glasser et al., 2016), and 8 subcortical regions defined bilaterally by the FreeSurfer atlas (Fischl, 2012; Fischl et al., 2002): thalamus, caudate, putamen, pallidum, hippocampus, amygdala, accumbens, and ventral diencephalon. This resulted in a 376-length vector for each of 5 regional qMT parameters for each participant.

### fMRI data acquisition and pre-processing

We used a multi-echo echoplanar imaging (EPI) sequence (Poser et al., 2006) to collect fMRI data under resting state conditions with the following parameters: TR = 2.57 s; echo times (TE_1,2,3_) = 15 ms, 34 ms and 54 ms; total acquisition time = 10 mins 42.5 s = 250 time points in each fMRI time series. Multi-echo EPI data were collected as 32 slices at −30 degrees downward pitch to the AC-PC line, with field of view 240 mm; matrix size 64×64; and voxel size:3.75 mm×3.75 mm×3.99 mm (for one site these parameters were marginally different, see SI page 3)

The first 6 volumes were discarded to ensure scanner equilibrium and the remaining data were pre-processed using multi-echo independent component analysis (ME-ICA; Kundu et al., 2012, 2013) to identify sources of variance in the fMRI time series that scaled linearly with TE and could thus be confidently regarded as BOLD signal. Other non-BOLD sources of variance, such as head movement, that do not scale with TE, were identified by ME-ICA and discarded. The retained independent components, representing BOLD contrast, were optimally recomposed to generate a broadband denoised fMRI time series at each voxel. This was bandpass filtered using the Maximal Overlap Discrete Wavelet Transform (“modwt” using “la8”, the Daubechies orthonormal compactly supported wavelet of length L=8), resulting in a BOLD signal oscillating in the frequency range 0.01-0.1 Hz (wavelet scales 2-4).

Geometric re-alignment was used to estimate 6 motion parameters for each participant (3 translation and 3 rotation parameters) which were used to calculate an overall estimate of motion – frame-wise displacement (FD) - defined as the Euclidean norm of motion and rotation derivatives in mm: 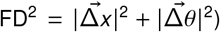. For each participant, mean FD was calculated by averaging the FD time series. A total of three scans were excluded due to high in-scanner motion hFDi_RMS_ > 0.3 mm or max(FD) > 1.3 mm and one subject was dropped due to excessively high mean correlation > 0.7.

Each pre-processed fMRI image was regionally parcellated into the same set of cortical and subcortical regions as the qMT data and the regional mean fMRI time series estimated for each cortical and sub-cortical region using the non-zero mean variant of the AFNI *3dROIstats* command (Cox, 1996). Thus we estimated a 376×244 regional time series matrix for each participant.

The functional connectivity between each regional pair of fMRI time series was estimated by Pearson’s correlation coefficient *r* for each possible pair of regions, resulting in a 376×376 symmetric association or functional connectivity matrix. The row (or column) means of this matrix comprise the vector of regional or nodal weighted degree (Fornito et al., 2016).

### Statistical analysis

We used Kolmgorov-Smirnov tests to assess between-group differences in whole brain distributions of regional PD, regional weighted degree, and functional connectivity or edge weight between each pair of regions. Between-group differences in PD and weighted degree were estimated and tested separately for all 376 brain regions. Likewise linear association between CRP and PD or weighted degree was estimated by regression for each regional node. Linear association between CRP and functional connectivity was estimated by regression for each of 70,500 edges in the whole brain connectome and for each of the 375 edges connecting each sub-cortical structure to all other nodes. All mass univariate significance tests of between-group differences in MRI metrics, or associations between CRP and MRI metrics, were controlled for multiple comparisons by the false discovery rate (P_FDR_ < 0.05).

